# Epidemiology of Aspergillosis Diagnoses in the U.S. using a National EHR Database, 2013-2023

**DOI:** 10.1101/2025.06.19.25329882

**Authors:** Brittany L. Morgan Bustamante, Elijah G. Martinez, Aidan Lee, Natalie J. Kane, Simon K. Camponuri, Rose M. Reynolds, Theo T. Snow, Juliana G.E. Bartels, Mark Hoffman, Theodore C. White, Justin V. Remais

## Abstract

**Importance:** Aspergillosis is an under-recognized fungal infection associated with rising hospitalizations, concerns about antifungal resistance, and substantial morbidity and mortality in the U.S. Yet, national data on aspergillosis remain fragmented due to absence of centralized surveillance.

**Objective:** To examine demographic and geographic variation and temporal trends in aspergillosis prevalence in the U.S. from 2013 to 2023; to assess how the COVID-19 pandemic influenced prevalence across population subgroups.

**Design:** Retrospective cohort study using a large, national electronic health record (EHR) database.

**Setting:** 142 healthcare systems in the U.S. using Oracle Health EHR systems, encompassing disparate care settings.

**Participants:** Adults aged 18 years and older who received care between January 1, 2013, and December 31, 2023. The cohort included over 76 million patients, and over 127 million person-years, with 20,764 aspergillosis diagnoses.

**Exposures:** Calendar year, patient characteristics (age, sex, race, ethnicity, residence type), and state of residence. COVID-19 was treated as a time-varying exposure.

**Main Outcomes and Measures:** Annual and overall prevalence of aspergillosis per 100,000 person-years; adjusted prevalence ratios (aPRs) by demographic group and state, estimated using quasi-Poisson and Bayesian spatiotemporal regression. COVID-19-related shifts in prevalence were assessed using estimated marginal means.

**Results:** Between 2013 and 2023, aspergillosis prevalence increased 5% annually, peaking in 2022. Rhode Island had the highest aPR across all years; Utah the lowest. Prevalence was higher in males (aPR: 1.37), older adults (aPR for ≥65 vs. 18 to 34: 4.95), and urban residents (rural aPR: 0.86). Post-COVID-19, prevalence increased disproportionately among Hispanic or Latino patients and several racial minority groups. Suggestive trends of increasing prevalence were also observed among rural residents.

**Conclusions and Relevance:** This study provides the most comprehensive national assessment to date of aspergillosis patterns in the U.S., revealing evidence of rising prevalence and emerging disparities shaped by the COVID-19 pandemic. Clinicians should consider regional and demographic risk patterns to support earlier diagnosis, guide antifungal treatment, and improve outcomes—especially among older adults and populations disproportionately affected by COVID-19. Clinicians practicing in rural or underserved areas should be particularly mindful of shifting risk profiles and advocate for improved diagnostic resources to reduce delays in care.

**Key Points:** 

**Question:** How has aspergillosis prevalence changed in the U.S. over time, and how did the COVID-19 pandemic influence prevalence among demographic and geographic subgroups?

**Findings:** In a nationally representative EHR cohort of over 76 million adults, aspergillosis prevalence increased by 5% annually from 2013 to 2023. After the emergence of COVID-19, prevalence increased disproportionately among Hispanic or Latino patients and racial minority groups.

**Meaning:** Findings suggest increasing aspergillosis prevalence in the U.S., with indications of widening disparities. While subject to data limitations, these trends underscore the importance of considering regional and demographic factors in clinical decision-making, particularly for populations disproportionately affected by COVID-19.

## Introduction

Aspergillosis is an infection caused by *Aspergillus* spp., a ubiquitous mold found indoors and outdoors.^1^ Over 24 *Aspergillus* spp. can infect humans, most commonly *A. fumigatus*, *A. terreu*s, and *A. flavus*,^2^ causing a range of syndromes depending on host immunity status.^3^ Most individuals inhale *Aspergillus* spores daily without consequence, but immunocompromised individuals or those with comorbidities can develop infections from mild to life-threatening.^4^ Invasive pulmonary and allergic aspergillosis pose serious risks to immunocompromised individuals, whereas immunocompetent individuals may experience less severe forms, such as developing aspergillomas.^5,6^

Despite the ubiquity of *Aspergillus*, aspergillosis epidemiology remains incomplete, partly due to variability in diagnosis and reporting.^7^ Global estimates suggest 250,000 cases of invasive aspergillosis and >3 million cases of pulmonary aspergillosis annually.^8,9^ In the U.S., the absence of centralized surveillance has made it difficult to assess national trends, even as aspergillosis-related hospitalizations increased by more than 44% between 2004 and 2013.^10^ Growing concerns about antifungal resistance,^11^ rising healthcare costs (over $670 million annually and $36,867 per hospitalization),^12^ and persistently high mortality—approximately 20% overall and nearly 100% if the infection becomes systemic^13,14^—underscore the need for more comprehensive monitoring and analysis.

Aspergillosis is more often diagnosed in urban patients^15^ and is one of the few fungal infections in which one disease state, pulmonary aspergillosis, is more common among higher-income individuals.^16^ Allergic forms of aspergillosis appear to be concentrated in the southern U.S., disproportionately affecting Black or African American populations.^17^ These patterns suggest potential influences from environmental exposures, structural inequities, and healthcare access. However, these factors remain underexplored, in part due to fragmented data. Without a comprehensive epidemiologic picture, it is difficult to disentangle the role of mediating and moderating factors in disease prevalence.

The COVID-19 pandemic heightened aspergillosis awareness, as COVID-19-associated pulmonary aspergillosis (CAPA) emerged as a severe complication among critically ill COVID patients.^12^ This brought renewed attention to the clinical significance of aspergillosis and underscored the importance of timely diagnosis and treatment.^19^ The pandemic also introduced substantial shifts in healthcare utilization—including changes in hospital admission patterns, care-seeking behavior, and diagnostic practices—that could have influenced the observed incidence of fungal infections. Understanding how these shifts may have affected aspergillosis diagnoses is critical for interpreting trends and preparing for future respiratory disease outbreaks.

A major challenge in assessing aspergillosis burden in the U.S. is the lack of surveillance.^20^ Existing estimates rely on administrative databases and electronic health records (EHRs), though prior studies have largely been limited to Medicare and Medicaid populations,^17,21^ inpatient versus outpatient comparisons,^22^ specific disease states,^10,23,24^ and state- or facility-specific assessments.^25–27^ This study aims to provide updated, population-wide estimates of aspergillosis diagnoses in the U.S. using a large, multi-state EHR dataset spanning various care settings.

By identifying geographic regions with rising case counts and estimating demographic differences in prevalence, this study offers insights into the spatial and social patterns of aspergillosis in the U.S. It also examines how the COVID-19 pandemic shaped diagnostic patterns, laying the foundation for understanding how social and structural dimensions of the pandemic—such as differential susceptibility, disrupted healthcare access, and uneven exposure—may have influenced disease burden. Together, these analyses contribute a comprehensive, population-wide perspective of this opportunistic infection’s evolving epidemiology in the U.S.

## Methods

### Study design

Data were extracted from Oracle EHR Real World Data (OERWD), a longitudinal, Health Insurance Portability and Accountability Act (HIPAA) compliant database covering over 76 million patients across 142 health systems. OERWD includes pharmacy, laboratory, admission, and billing data from the EHR of health systems that have a data use agreement with Oracle.^28^ All data are date and time stamped, providing a temporal relationship between treatment patterns and clinical information. The University of California, Berkeley ethics committee deemed this study non-human subjects research.

We created a retrospective cohort by querying OERWD in Oracle’s HealtheDataLab cloud-based platform for all patients who had at least one healthcare encounter with any recorded diagnosis each year from January 1, 2013-December 31, 2023. Patient totals were stratified by self-identified sex/gender (male or female), race (American Indian/Alaskan Native, Black or African American, Asian/Native Hawaiian or Pacific Islander, White, and some other race), ethnicity (Hispanic or Latino and non-Hispanic or Latino), residence type (urban or rural), age (18-34, 35-64, ≥65 years), and state of residence. Cases were defined as patients with an International Classification of Diseases (ICD) 9^th^ or 10^th^ revision code indicative of an aspergillosis diagnosis (**Table S1**) listed in any encounter record position. Patients could have multiple diagnoses over the study period, given the potential for repeat infections. However, patients with more than one aspergillosis diagnosis within a one-year period, possibly reflecting repeat visits for the same infection, were treated as a single case.

### Statistical Analysis

To adjust for differential subgroup representation in OERWD relative to the U.S. population, we calculated nationally-representative population-based aspergillosis prevalence estimates using post-stratification weighting.^29^ To generate population-based estimates, the OERWD cohort was aggregated to match the American Community Survey (ACS) strata (year, sex/gender, race/ethnicity, age group, and state),^30^ and post-stratified weights (**Equation S1**) were applied to adjust for demographic differences.^31^ Weights were based on ACS 5-year population estimates (2013-2023), and population-based prevalence estimates were calculated using weighted case counts and cohort totals.

To examine geographic and temporal patterns in the continental U.S., we implemented a Bayesian hierarchical spatiotemporal model using Integrated Nested Laplace Approximation (INLA).^32^ Weighted annual case counts per state were modeled using Poisson regression. Spatial dependence was accounted for using a Besag-York-Mollié model, which incorporates structured and unstructured spatial random effects based on a state boundary adjacency matrix. This modeling approach helps account for unmeasured area-level factors and spatial autocorrelation in prevalence that may arise from geographic variation in unmeasured confounders. Temporal trends were modeled using a first-order random walk latent effect, allowing for smooth year-to-year variation. State-specific random slopes captured differential temporal patterns across states. The model was adjusted for sex, race/ethnicity, and age group. Expected cases in each state, computed via indirect standardization using national age, year, and race/ethnicity stratified rates in the *SpatialEpi* package,^33^ were included as an offset and served as the reference. State-specific adjusted prevalence ratios (aPR) and 95% credible intervals (CrIs) were estimated relative to the national average.

To compare prevalence across demographic groups, we fit a multivariable quasi-Poisson model with the log of the OERWD cohort as an offset. Because post-stratification weights could not be calculated for residence type strata, we used robust standard errors to generate population-based estimates.^34^ aPRs and 95% confidence intervals (CI) were estimated from a model including fixed effects for sex, race, ethnicity, residence type, age group, state, and calendar year. To assess COVID-19’s impact on prevalence trends, we centered year at 2019 and included interaction terms between a binary COVID-19 indicator (0=pre-COVID-19, 1=COVID-19 years) and race, ethnicity, and residence type to examine subgroup-specific changes, along with a COVID-by-year interaction to adjust for shifts in temporal trends during the pandemic. Post hoc analyses using the *emmeans* package^35^ estimated the changes in prevalence (post-vs. pre-COVID-19) by demographic and residence group. Estimated marginal means were conditioned on the centered year (i.e., 2019) to isolate the step change associated pandemic onset, adjusting for pre-pandemic temporal trends. All contrasts were adjusted for multiple comparisons using the Sidak method.^36^

## Results

### OERWD Cohort and Aspergillosis Patient Characteristics

The OERWD cohort included 76,192,643 patients and 127,414,388 person-years over the 11-year study period (**Table 1**). Most of the cohort self-identified as White (73.7%), non-Hispanic or Latino (75.9%), female (58.5%), aged 35 to 64 years (46.8%), and resided in urban areas (81.2%; **Table 1**). A total of 20,764 aspergillosis cases were identified in the cohort.

**Table 1:** Number and percent of patients included in the Oracle EHR Real World Data (OERWD) cohort and diagnosed with aspergillosis, by patient characteristics–United States, 2013-2023.

| Characteristic | OERWD Cohort<br>Person-Years (%) | Aspergillosis Cases<br>N (%) |
| --- | --- | --- |
| N | 127,414,388 | 20,764 |
| **Sex** |  |  |
| Male | 52,790,651 (41.4) | 10,486 (50.5) |
| Female | 74,579,070 (58.5) | 10,268 (49.5) |
| Other or unknown | 44,667 (<1) | 10 (<1) |
| **Race** |  |  |
| White | 93,934,750 (73.7) | 15,329 (73.8) |
| Black or African American | 11,944,842 (9.4) | 1,781 (8.6) |
| American Indian/Alaska Native | 974,719 (<1) | 134 (<1) |
| Asian/ Pacific Islander | 3,194,555 (2.5) | 702 (3.4) |
| Some Other Race | 10,195,552 (8.0) | 2,176 (10.5) |
| Unknown | 7,169,970 (5.6) | 642 (3.1) |
| **Ethnicity** |  |  |
| Hispanic or Latino | 19,964,215 (15.7) | 3,207 (15.5) |
| Non-Hispanic or Latino | 96,752,537 (75.9) | 15,966 (76.9) |
| Unknown | 10,697,636 (8.4) | 1,591 (7.7) |
| **Age Group** |  |  |
| 18 to 34 years | 32,127,328 (25.2) | 2,031 (9.8) |
| 35 to 64 years | 59,634,609 (46.8) | 8,781 (42.3) |
| 65 years and over | 35,652,451 (27.9) | 9,952 (47.9) |
| **Residence Type** |  |  |
| Urban residence | 103,423,958 (81.2) | 17,382 (83.7) |
| Rural residence | 22,728,960 (17.8) | 3,271 (15.8) |
| Unknown | 1,261,470 (<1) | 111 (<1) |

Aspergillosis cases were more likely to be male (50.5%) compared to the cohort (41.4%) and were older, with nearly half aged 65 years and over (47.9%) versus 27.9% in the cohort (**Table 1**). While the racial distribution was similar, Asian/Pacific Islander individuals were slightly overrepresented (3.4% in the cases and 2.5% in cohort), as were individuals identifying as Some Other Race (10.5% in the cases and 8.0% in the cohort; **Table 1**). Urban residence was more common among aspergillosis cases (83.7%) than the cohort (81.2%; **Table 1**). Geographically, the highest percentage of cases were reported in California (23.9%), and lowest was reported in Louisiana (<1%; **Figure S1**).

### Aspergillosis Prevalence and Spatiotemporal Trends

The population-based aspergillosis prevalence among U.S. adults aged 18 years and over from 2013 to 2023 was 15.26 diagnoses per 100,000 person-years. Prevalence peaked in 2022 at 18.04 diagnoses per 100,000 person-years, with the lowest recorded prevalence in 2013 (11.99 diagnoses per 100,000 person-years) (**Figure S2**). Prevalence was highest in Rhode Island and Arizona, and lowest in Utah and Nevada (**Figure 1A**). There was substantial variation in aspergillosis across the continental U.S. Rhode Island’s prevalence, while decreasing over the study period, substantially exceeded the national average each year, ranging from an aPR of 11.04 (95% CrI: 6.74-17.09) in 2013 to 6.20 (95% CrI: 3.75-9.59) in 2023 (**Figure 1B**), indicating an observed prevalence six to 11 times higher than expected. In contrast, Utah remained well below the national average, with aPR decreasing slightly from 0.20 (95% CrI: 0.09-0.38) in 2013 to 0.17 (95% CrI: 0.07-0.35) in 2023 (**Figure 1B**). Alabama experienced the largest aPR increase from 0.31 (95% CrI: 0.20-0.46) in 2013 to 1.42 (95% CrI: 1.14-1.72) in 2023 (**Figure 1B**). California exhibited a steady upward trend, increasing from an aPR of 1.00 (95% CrI: 0.92-1.09) in 2013 to 1.81 (95% CrI: 1.71-1.91) in 2023 (**Table S2**). Several other states—including Delaware, Kentucky, Missouri, Nebraska, New Jersey, New Mexico, and South Dakota—experienced significant increases in prevalence over the study period (**Table S2**). While the magnitude varied, most states in the analysis experienced increasing aPRs over the study period, peaking between 2020 and 2022 (**Figure 2**).

**Figure 1:**
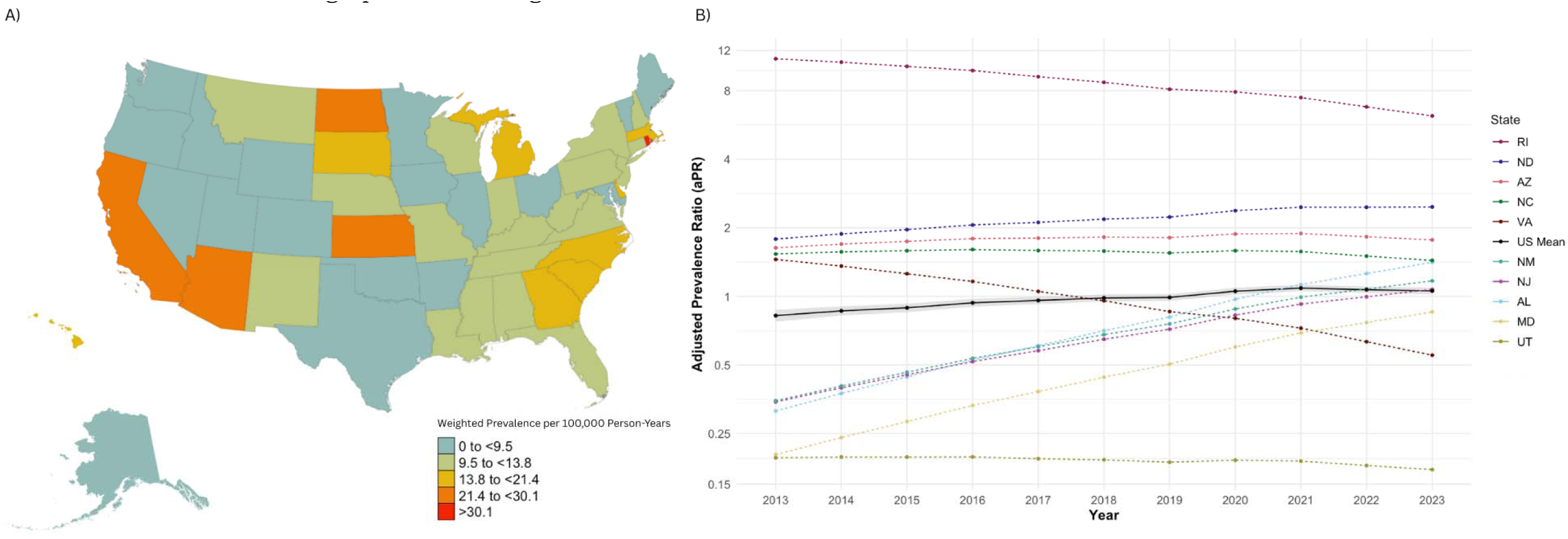
(A)–Population-based prevalence of aspergillosis diagnoses per 100,000 person-years across the study period (2013-2023), by state; (B)–Annual adjusted prevalence ratio (aPR) for the five states with the highest and lowest initial aPRs compared to the national average. Weighted prevalence and aPRs calculated using post-stratified weighted case and cohort counts; Panel B – the legend is ordered to match the order of the lines on the graph at 2013; the black line represents the national average aPR and grey band represents the national average 95% credible interval; the reference for the aPRs is the expected case counts computed via indirect standardization; graph is on the logscale.

**Figure 2:**
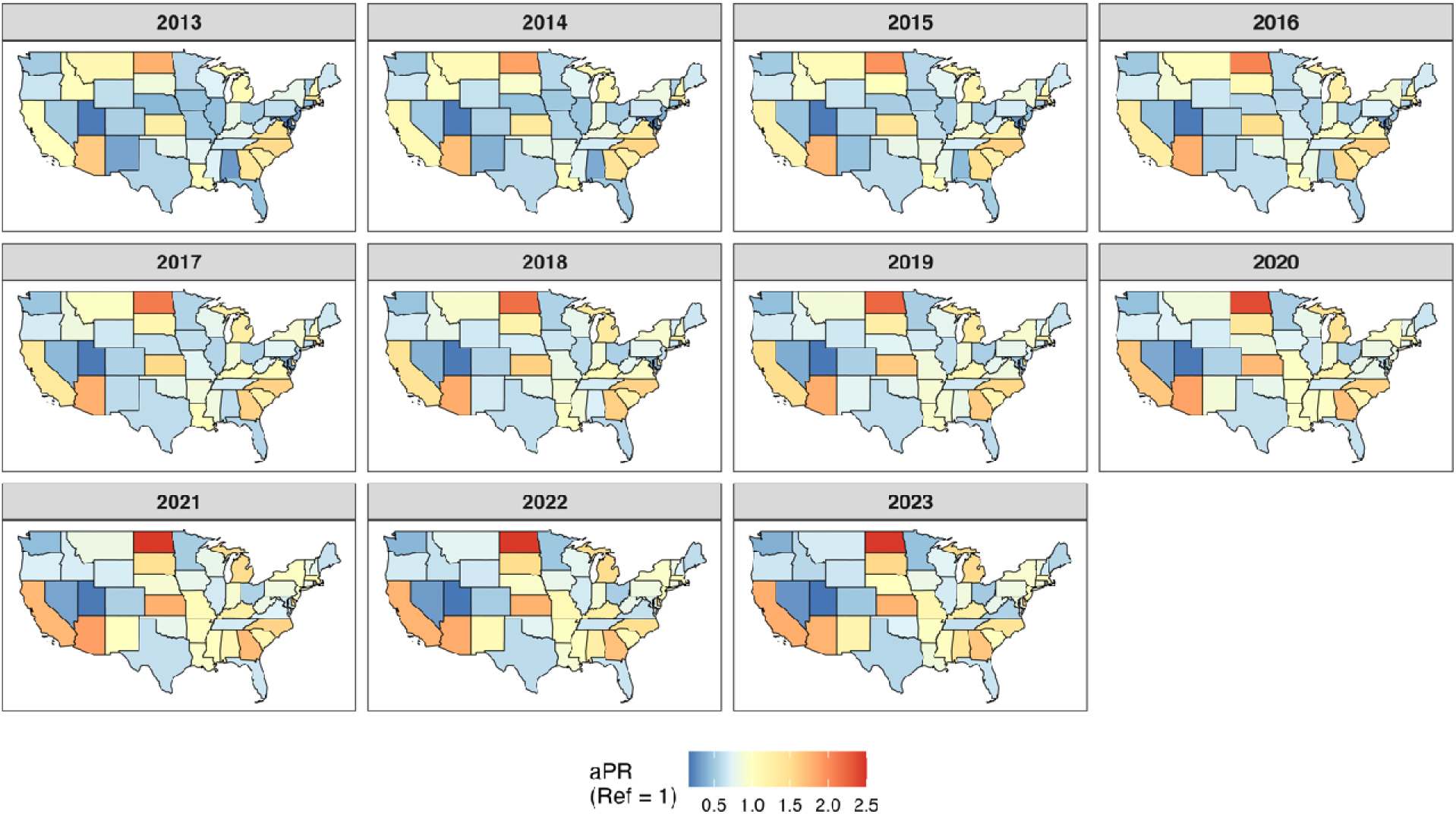
State-level adjusted prevalence ratios (aPR) for aspergillosis diagnoses in U.S. adults. Annual estimates from weighted Oracle EHR Real World Data, 2013 to 2023. aPR adjusted for sex/gender, age group, race and ethnicity; aPR calculated using post-stratification weighted case and cohort counts and the expected number of diagnoses in each state as the offset/reference; Analysis only included continental U.S. due to contiguity-based spatial weights requiring at least one neighbor.

### Aspergillosis Prevalence by Sex, Race, Ethnicity, Age, and Residence Type

A total of 111,203,140 person-years in the cohort (87.3%) had complete demographic data and were included in the multivariable analysis. Over the study period, prevalence was significantly higher in males compared to females (aPR: 1.37; 95% CI: 1.32-1.42; *P*<.001) and among patients aged 65 years relative to those aged 18 to 34 years (aPR: 4.95; 95% CI: 4.65-5.28; *P*<.001; **Table 2**). Few differences were seen by racial identity. Prevalence among American Indian/Alaska Native and Asian/Pacific Islander patients was not significantly different from White patients (**Table 2**). Patients identifying as Black or African American and “Some Other Race” had slightly higher aspergillosis prevalence compared to White patients (aPR: 1.08; 95% CI: 1.01-1.16; *P*=0.03 and aPR: 1.25; 95% CI: 1.16-1.33; *P*<.001, respectively, **Table 2**).

**Table 2:** Multivariable quasi-Poisson regression results: association of demographic characteristics and COVID-19 with aspergillosis prevalence–United States, 2013-2023.

| Variable | Adjusted Prevalence |  |  |
| --- | --- | --- | --- |
|  | Ratio | Robust 95% CI | P-value |
| (Intercept) | 0.00 | (0.00, 0.00) | <0.001 |
| White | *ref* | *ref* | *ref* |
| American Indian/Alaska Native | 0.78 | (0.58, 1.05) | 0.11 |
| Asian/Pacific Islander | 0.93 | (0.82, 1.06) | 0.29 |
| Black or African American | 1.06 | (0.97, 1.16) | 0.22 |
| Some Other Race | 1.25 | (1.14, 1.37) | <0.001 |
| Hispanic or Latino* | 0.84 | (0.78, 0.92) | <0.001 |
| Rural | 0.86 | (0.80, 0.92) | <0.001 |
| Male** | 1.37 | (1.32, 1.42) | <0.001 |
| Age 18 to 34 | *ref* | *ref* | *ref* |
| Age 35 to 64 | 2.56 | (2.40, 2.73) | <0.001 |
| Age 65 and over | 4.95 | (4.65, 5.28) | <0.001 |
| Year*** | 1.05 | (1.04, 1.07) | <0.001 |
| COVID-19<sup>+</sup> | 1.02 | (0.94, 1.10) | 0.68 |
| Year*COVID-19 | 0.95 | (0.92, 0.98) | <0.001 |

*Note: Model controlled for state of residence; Robust SE (standard errors) calculated using sandwich estimator; ref = reference group; CI = confidence interval*
*\*Reference is non-Hispanic or Latino*
*\*\*Reference is female sex/gender*
*\*\*\*Year is a continuous variable centered at 2019*
*<sup>+</sup>Reference is pre-COVID-19 period (2013-2019)*

Hispanic or Latino patients were less likely to be diagnosed with aspergillosis than non-Hispanic or Latino patients (aPR: 0.92; 95% CI: 0.87-0.99; *P*=0.01), as were rural residents when compared to those living in urban areas (aPR: 0.89; 95% CI: 0.84-0.93; *P*<.001; **Table 2**).

### Aspergillosis Prevalence and the COVID-19 Pandemic

Before the COVID-19 pandemic, aspergillosis prevalence increased steadily by approximately 5.3% per year (95% CI: 4.0%-6.9%; *P*<.001; **Table 2**). Following the emergence of COVID-19, the annual increase in aspergillosis prevalence stabilized, with the trend decreasing by 5.2% (95% CI: 2.6%-7.7%; *P<.*001; **Table 2**) during the post-COVID period compared to the pre-COVID period. Accounting for this shift in trend, post-hoc analyses revealed disproportionate increases in certain demographic subgroups (**Table S3**). Within all racial groups, Hispanic or Latino patients experienced a larger relative increase in prevalence post-COVID-19 compared to pre-COVID-19 (aPR: 1.36; 95% CI: 1.15-1.61) than did non-Hispanic or Latino patients (aPR: 1.15; 95% CI: 0.99-1.34; **Figure 3A**), after adjusting for age, sex, year, residence type, and state of residence. Rural residents of all races also had a slightly greater increase in prevalence post-COVID-19 relative to the pre-pandemic period (aPR: 1.29; 95% CI: 1.09-1.53) than urban residents (aPR: 1.22; 95% CI: 1.05-1.42; **Figure 3B**). Patients identifying as Asian/Pacific Islander, American Indian/Alaska Native, and Hispanic or Latino (any race) experienced significantly greater post-COVID-19 increases in prevalence compared to White and non-Hispanic or Latino patients, with relative increases of 32% (95% CI: 26%-39%; *P*<.001), 18% (95% CI: 11%-24%; *P*<.001), and 21% (95% CI: 16%-25%; *P*<.001), respectively (**Table S4**).

**Figure 3:**
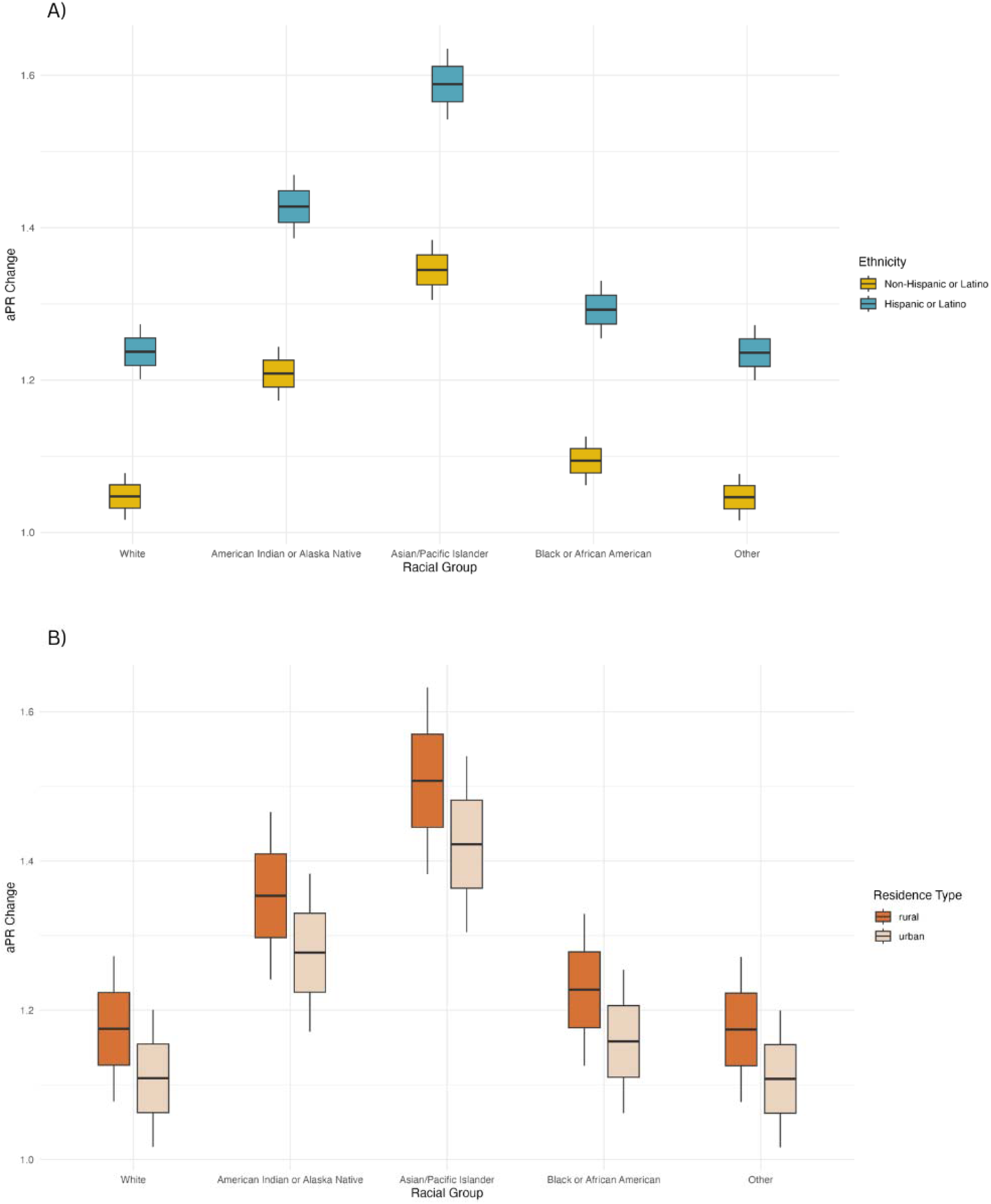
Adjusted prevalence ratios (aPR) for aspergillosis from pre- to post-emergence of COVID-19 across ethnicity (A) and residence type (B) for all racial groups–United States, 2013 to 2023. Panels display median prevalence ratios with interquartile (IQR) ranges and 1.5*IQR whiskers; ethnicity estimates are averaged over residence type and residence type estimates are averaged over ethnicity groups. The pre-COVID-19 period spans 2013-2019, and the post-COVID-19 period spans 2020-2023.

## Discussion

Our study provides a comprehensive analysis of aspergillosis trends in the U.S. over an 11-year period, leveraging a large, diverse cohort of patients weighted to generate nationally representative prevalence estimates. Results suggest an overall increase in aspergillosis prevalence between 2013 and 2023, with the peak observed in 2021. Previous research has documented both increasing and decreasing trends of *Aspergillus-*associated infections in the U.S.^17,22,23,37^ However, these studies were limited to specific forms of the disease, focused solely on inpatient populations, or restricted to high-risk groups. Our work builds upon these efforts by providing a broader evaluation of aspergillosis prevalence across a longer temporal span, diverse healthcare settings, demographic subgroups, and all-payer types, while applying national weighting to achieve greater representativeness.

Consistent with prior research,^24,38^ non-Hispanic or Latino White patients were more likely to be aspergillosis cases than other racial and ethnic groups. This pattern may reflect differences in underlying conditions that predispose individuals to aspergillosis, such as transplantation, pulmonary disease, and immunosuppressive therapies, which are higher among non-Hispanic or Latino White individuals.^14,39^ Aspergillosis was also more common in urban than rural patients, which could be related to socioeconomic factors. Previous studies have shown that aspergillosis is more prevalent in individuals from higher-income areas^16^—populations that are more often concentrated in urban regions—possibly due to greater access to specialized healthcare, increased likelihood of receiving a transplant, and better management of chronic conditions like cystic fibrosis.^15,37^

From 2013 to 2019, aspergillosis prevalence appeared to increase steadily, with trends persisting after adjustment for demographic and geographic factors. After the onset of the COVID-19 pandemic in 2020, this trend in prevalence significantly slowed. The reasons for the pre-pandemic increase remain unclear but may reflect changes in underlying host risk (e.g., rising use of immunosuppressive therapies, aging population with more at-risk patients),^10^ diagnostic practices,^40^ or heightened clinical awareness.^41^ In contrast, the decreasing post-pandemic trend may be attributable to changes in healthcare utilization,^42^ shifts in healthcare priorities,^43^ public health interventions (e.g., masking, reduced travel),^44^ or a possible "saturation effect" (i.e., high-risk populations captured early in the pandemic).^45^

To the best of our knowledge, this is the first study to identify COVID-19 pandemic-associated shifts in aspergillosis prevalence patterns. We observed notable shifts in demographic groups most likely to be diagnosed with aspergillosis following the emergence of COVID-19, with increased prevalence among racial and ethnic minority populations and rural residents. We identified disproportionate increases among Hispanic or Latino patients compared to non-Hispanic or Latino, of all races, as well as suggestive increased prevalence among rural residents compared to urban residents. These disparities may reflect differences in baseline health status, access to timely diagnosis and treatment, or the disproportionate impact of severe COVID-19 in these populations.^46,47^ Hispanic and rural patients may have faced greater exposure risks or delayed healthcare access, compounding the observed trends.^48^ Further, the higher post-COVID-19 prevalence observed among Asian/Pacific Islander and American Indian/Alaska Native patients compared to White patients highlights the importance of considering structural and social determinants of health in future studies.^49,50^ Evidence suggests that conventional risk factors for invasive aspergillosis are not common among individuals with CAPA,^51^ suggesting factors specifically related to the COVID-19 pandemic such as disruptions in healthcare access, differential rates of severe disease, and variations in hospitalization and intensive care unit admissions could contribute to these disparities. However, further research is needed to elucidate the mechanisms underlying these patterns and to identify targeted interventions to mitigate risk.

Our findings also represent the first comprehensive assessment of the spatial and temporal patterns in all-cause aspergillosis prevalence at the state level across the continental U.S. After adjusting for patient demographics, we observed substantial heterogeneity in state-level prevalence. Rhode Island consistently had the highest aPR throughout the study period, which may reflect a true increase in disease prevalence given its population structure aligns with groups identified in the literature as being at higher risk. However, this may also reflect uneven healthcare system coverage in the dataset. Although post-stratification weights were applied to enhance representativeness, residual bias may persist due to unmeasured factors not included in our weighting scheme such as chronic disease burden, diagnostic practices, or healthcare-seeking behavior. Notably, several other states – including Alabama, Delaware, Kentucky, Missouri, Nebraska, New Jersey, New Mexico, and South Dakota – experienced statistically significant increases in prevalence over time. These findings underscore the need for research on how healthcare access, socioeconomic conditions, and urbanization shape aspergillosis distribution.^16,52–54^

Despite its strengths, this study has several limitations. As with all analyses using EHR data, variability in diagnostic coding practices, case definitions, patient populations, and geographic representation may introduce misclassification or bias. To mitigate these risks, we created post-stratification weights to enhance generalizability and conducted data quality checks, including excluding implausible records and using diagnosis-linked encounters to generate patient counts for weighting. These steps help address possible biases related to data flows or health system-specific factors affecting data completeness.^55^ Another key limitation is the absence of laboratory confirmation for aspergillosis diagnoses. Case definitions relied on ICD codes, which are limited in granularity and subject to potential coding inaccuracies.^56^ Previous studies have estimated the sensitivity of ICD codes for aspergillosis to be 84%, with a positive predictive value ranging from 65% to 67%.^57,58^ While future studies should aim to replicate similar analyses among culture-confirmed cases, culturing rates in the U.S. remain low.^58^ As such, a comprehensive understanding of the disease’s epidemiology will require integrating findings from both clinical and microbiologically confirmed datasets. Finally, because aspergillosis is not a nationally reportable disease, no surveillance system or gold standard currently exists to validate our estimates.

These findings provide one of the most comprehensive, nationally representative assessments to date of aspergillosis prevalence across the U.S., highlighting important temporal, geographic, and demographic patterns. We documented an overall increase in aspergillosis prevalence over the past decade, peaking in 2021, with shifts in groups most affected during the COVID-19 pandemic across race, ethnicity, and geography. These findings underscore the need to understand how social, structural, and clinical factors interact to shape aspergillosis disease risk– particularly in the context of respiratory-illness public health crises.

## Data Availability

Due to proprietary restrictions, the datasets analyzed in this study are not publicly available. Researchers from academia, healthcare systems, or life sciences organizations can access OERWD if their institution contributes de-identified data to the dataset or partners with Oracle Health through the Learning Health Network to use HealtheDataLab, a cloud-based distributed learning platform for approved research projects. Other researchers or organizations may apply for access to OERWD, subject to approval. Analytic code for this project can be found at: https://github.com/b-lmb/Aspergillosis-epidemiology

## Data Availability

Due to proprietary restrictions, the datasets analyzed in this study are not publicly available.
Researchers from academia, healthcare systems, or life sciences organizations can access OERWD if their institution contributes de-identified data to the dataset or partners with Oracle Health through the Learning Health Network to use HealtheDataLab, a cloud-based distributed learning platform for approved research projects. Other researchers or organizations may apply for access to OERWD, subject to approval.

