## Supplemental Documents for "Epidemiology of Aspergillosis Diagnoses in the U.S. using a National EHR Database, 2013-2023"

**Table S1. ICD-9 and ICD-10 codes used to identify aspergillosis cases**

|  | **ICD-9** | | **ICD-10** |
| --- | --- | --- | --- |
| **Disease** | **Diagnosis code** | **Manifestation Code** | **Diagnosis Code** |
| Aspergillosis | 117.3 | 484.6, 518.6.78 | B44* |

**Table S2: Adjusted prevalence ratio (aPR) and 95% credible interval estimates for aspergillosis by state and year using population-weighted data from Oracle EHR Real World Data (OERWD)–United States, 2013-2023**

| **State** | **Year** | | | | | | | | | | |
| --- | --- | --- | --- | --- | --- | --- | --- | --- | --- | --- | --- |
|  | **2013** | **2014** | **2015** | **2016** | **2017** | **2018** | **2019** | **2020** | **2021** | **2022** | **2023** |
| **AL** | 0.31  (0.20, 0.46) | 0.37  (0.26, 0.53) | 0.44  (0.32, 0.60) | 0.52  (0.40, 0.68) | 0.61  (0.48, 0.76) | 0.71  (0.58, 0.85) | 0.81  (0.68, 0.95) | 0.97  (0.83, 1.13) | 1.13  (0.96, 1.31) | 1.26  (1.05, 1.49) | 1.41  (1.14, 1.72) |
| **AR** | 0.75  (0.35, 1.40) | 0.78  (0.40, 1.40) | 0.82  (0.44, 1.39) | 0.85  (0.48, 1.40) | 0.87  (0.51, 1.39) | 0.89  (0.53, 1.41) | 0.90  (0.53, 1.44) | 0.96  (0.55, 1.55) | 0.99  (0.55, 1.64) | 0.98  (0.52, 1.69) | 0.98  (0.48, 1.76) |
| **AZ** | 1.63  (1.48, 1.80) | 1.70  (1.56, 1.84) | 1.74  (1.63, 1.87) | 1.79  (1.69, 1.91) | 1.80  (1.71, 1.90) | 1.82  (1.73, 1.91) | 1.81  (1.73, 1.90) | 1.88  (1.79, 1.97) | 1.89  (1.80, 1.99) | 1.83  (1.73, 1.93) | 1.77  (1.65, 1.89) |
| **CA** | 1.00  (0.92, 1.09) | 1.10  (1.02, 1.17) | 1.18  (1.12, 1.26) | 1.28  (1.21, 1.35) | 1.36  (1.29, 1.42) | 1.44  (1.38, 1.50) | 1.51  (1.44, 1.57) | 1.65  (1.58, 1.72) | 1.74  (1.67, 1.82) | 1.77  (1.69, 1.86) | 1.81  (1.71, 1.91) |
| **CO** | 0.54  (0.38, 0.73) | 0.55  (0.41, 0.72) | 0.55  (0.43, 0.70) | 0.56  (0.45, 0.68) | 0.55  (0.46, 0.66) | 0.55  (0.46, 0.65) | 0.54  (0.45, 0.63) | 0.55  (0.45, 0.66) | 0.54  (0.44, 0.67) | 0.52  (0.40, 0.66) | 0.50  (0.37, 0.65) |
| **CT** | 0.44  (0.32, 0.59) | 0.49  (0.37, 0.63) | 0.53  (0.42, 0.67) | 0.58  (0.47, 0.71) | 0.62  (0.53, 0.73) | 0.67  (0.58, 0.77) | 0.71  (0.63, 0.80) | 0.79  (0.70, 0.88) | 0.84  (0.74, 0.95) | 0.87  (0.75, 1.00) | 0.90  (0.76, 1.06) |
| **DE** | 0.83  (0.65, 1.05) | 0.92  (0.74, 1.12) | 1.00  (0.84, 1.19) | 1.09  (0.94, 1.27) | 1.17  (1.03, 1.32) | 1.25  (1.12, 1.39) | 1.32  (1.20, 1.46) | 1.46  (1.32, 1.60) | 1.56  (1.40, 1.73) | 1.60  (1.41, 1.80) | 1.65  (1.42, 1.90) |
| **FL** | 0.47  (0.38, 0.56) | 0.50  (0.42, 0.58) | 0.52  (0.45, 0.60) | 0.55  (0.49, 0.62) | 0.57  (0.51, 0.63) | 0.59  (0.54, 0.64) | 0.60  (0.55, 0.65) | 0.64  (0.59, 0.69) | 0.66  (0.60, 0.71) | 0.65  (0.59, 0.72) | 0.65  (0.58, 0.73) |
| **GA** | 1.37  (1.06, 1.74) | 1.44  (1.15, 1.77) | 1.49  (1.23, 1.79) | 1.55  (1.32, 1.81) | 1.57  (1.37, 1.80) | 1.61  (1.41, 1.82) | 1.62  (1.43, 1.82) | 1.70  (1.49, 1.92) | 1.72  (1.49, 1.98) | 1.69  (1.43, 1.98) | 1.66  (1.36, 1.99) |
| **IA** | 0.46  (0.35, 0.60) | 0.50  (0.39, 0.63) | 0.54  (0.44, 0.66) | 0.58  (0.49, 0.69) | 0.61  (0.53, 0.71) | 0.65  (0.57, 0.74) | 0.68  (0.60, 0.77) | 0.74  (0.65, 0.85) | 0.79  (0.67, 0.91) | 0.80  (0.67, 0.95) | 0.82  (0.66, 1.00) |
| **ID** | 1.01  (0.49, 1.89) | 0.98  (0.51, 1.72) | 0.94  (0.52, 1.57) | 0.90  (0.53, 1.44) | 0.85  (0.52, 1.32) | 0.80  (0.50, 1.23) | 0.75  (0.47, 1.15) | 0.73  (0.45, 1.14) | 0.70  (0.41, 1.11) | 0.64  (0.35, 1.05) | 0.58  (0.30, 1.01) |
| **IL** | 0.45  (0.34, 0.60) | 0.49  (0.38, 0.62) | 0.52  (0.42, 0.63) | 0.55  (0.45, 0.65) | 0.57  (0.48, 0.66) | 0.59  (0.51, 0.68) | 0.61  (0.53, 0.69) | 0.65  (0.57, 0.75) | 0.68  (0.58, 0.79) | 0.68  (0.57, 0.81) | 0.68  (0.55, 0.83) |
| **IN** | 0.81  (0.69, 0.94) | 0.84  (0.74, 0.95) | 0.86  (0.77, 0.96) | 0.88  (0.80, 0.97) | 0.88  (0.81, 0.96) | 0.89  (0.83, 0.96) | 0.88  (0.83, 0.94) | 0.91  (0.86, 0.98) | 0.92  (0.85, 0.98) | 0.88  (0.81, 0.96) | 0.85  (0.78, 0.94) |
| **KS** | 1.26  (0.99, 1.59) | 1.34  (1.09, 1.64) | 1.42  (1.18, 1.69) | 1.49  (1.28, 1.73) | 1.54  (1.36, 1.74) | 1.60  (1.43, 1.77) | 1.63  (1.48, 1.79) | 1.74  (1.59, 1.90) | 1.79  (1.62, 1.97) | 1.78  (1.59, 1.99) | 1.77  (1.55, 2.02) |
| **KY** | 0.78  (0.63, 0.97) | 0.85  (0.70, 1.02) | 0.90  (0.77, 1.06) | 0.97  (0.84, 1.11) | 1.01  (0.89, 1.14) | 1.06  (0.94, 1.19) | 1.10  (0.98, 1.24) | 1.19  (1.04, 1.35) | 1.25  (1.08, 1.44) | 1.26  (1.06, 1.48) | 1.27  (1.04, 1.53) |
| **LA** | 1.03  (0.40, 2.24) | 1.03  (0.42, 2.17) | 1.03  (0.43, 2.10) | 1.02  (0.43, 2.06) | 0.99  (0.42, 2.00) | 0.98  (0.41, 1.98) | 0.95  (0.39, 1.96) | 0.96  (0.37, 2.05) | 0.95  (0.35, 2.09) | 0.90  (0.31, 2.07) | 0.86  (0.27, 2.07) |
| **MA** | 1.35  (1.17, 1.54) | 1.35  (1.20, 1.52) | 1.34  (1.21, 1.48) | 1.33  (1.21, 1.45) | 1.29  (1.19, 1.39) | 1.25  (1.16, 1.35) | 1.20  (1.11, 1.29) | 1.20  (1.10, 1.31) | 1.16  (1.06, 1.28) | 1.09  (0.97, 1.21) | 1.01  (0.89, 1.15) |
| **MD** | 0.20  (0.14, 0.28) | 0.24  (0.18, 0.31) | 0.28  (0.22, 0.36) | 0.33  (0.27, 0.41) | 0.38  (0.32, 0.45) | 0.44  (0.38, 0.51) | 0.50  (0.44, 0.57) | 0.60  (0.52, 0.68) | 0.69  (0.59, 0.80) | 0.77  (0.64, 0.91) | 0.85  (0.69, 1.04) |
| **ME** | 0.66  (0.53, 0.82) | 0.67  (0.55, 0.81) | 0.67  (0.57, 0.79) | 0.68  (0.59, 0.78) | 0.67  (0.59, 0.75) | 0.66  (0.58, 0.74) | 0.64  (0.57, 0.72) | 0.65  (0.57, 0.74) | 0.64  (0.55, 0.74) | 0.61  (0.51, 0.72) | 0.57  (0.47, 0.70) |
| **MI** | 1.05  (0.54, 1.85) | 1.12  (0.62, 1.86) | 1.17  (0.70, 1.86) | 1.24  (0.78, 1.87) | 1.28  (0.84, 1.86) | 1.33  (0.90, 1.89) | 1.36  (0.94, 1.91) | 1.46  (1.00, 2.06) | 1.51  (1.01, 2.19) | 1.52  (0.97, 2.26) | 1.52  (0.93, 2.37) |
| **MN** | 0.58  (0.41, 0.79) | 0.58  (0.43, 0.76) | 0.58  (0.45, 0.73) | 0.58  (0.46, 0.71) | 0.56  (0.46, 0.68) | 0.55  (0.45, 0.67) | 0.53  (0.43, 0.65) | 0.54  (0.42, 0.67) | 0.52  (0.40, 0.68) | 0.49  (0.36, 0.66) | 0.46  (0.32, 0.65) |
| **MO** | 0.50  (0.43, 0.58) | 0.56  (0.49, 0.64) | 0.62  (0.55, 0.69) | 0.69  (0.62, 0.76) | 0.74  (0.69, 0.81) | 0.81  (0.75, 0.87) | 0.87  (0.81, 0.93) | 0.97  (0.91, 1.04) | 1.05  (0.98, 1.13) | 1.10  (1.02, 1.19) | 1.15  (1.05, 1.26) |
| **MS** | 0.71  (0.41, 1.15) | 0.76  (0.47, 1.16) | 0.79  (0.52, 1.15) | 0.83  (0.58, 1.15) | 0.85  (0.63, 1.13) | 0.88  (0.68, 1.13) | 0.90  (0.71, 1.13) | 0.96  (0.75, 1.21) | 0.99  (0.76, 1.27) | 0.99  (0.73, 1.31) | 0.98  (0.69, 1.36) |
| **MT** | 1.11  (0.88, 1.37) | 1.09  (0.90, 1.31) | 1.06  (0.89, 1.24) | 1.03  (0.89, 1.18) | 0.98  (0.86, 1.10) | 0.93  (0.83, 1.05) | 0.88  (0.77, 0.99) | 0.86  (0.75, 0.98) | 0.82  (0.70, 0.96) | 0.75  (0.62, 0.90) | 0.69  (0.56, 0.85) |
| **NC** | 1.54  (1.14, 2.02) | 1.57  (1.21, 2.00) | 1.59  (1.27, 1.96) | 1.61  (1.32, 1.93) | 1.59  (1.34, 1.87) | 1.58  (1.35, 1.84) | 1.55  (1.32, 1.81) | 1.59  (1.34, 1.87) | 1.57  (1.30, 1.89) | 1.50  (1.21, 1.85) | 1.44  (1.11, 1.83) |
| **ND** | 1.78  (0.81, 3.46) | 1.88  (0.90, 3.50) | 1.96  (0.99, 3.53) | 2.06  (1.07, 3.60) | 2.11  (1.12, 3.65) | 2.18  (1.16, 3.77) | 2.23  (1.16, 3.90) | 2.38  (1.20, 4.26) | 2.46  (1.18, 4.57) | 2.46  (1.11, 4.76) | 2.47  (1.04, 5.01) |
| **NE** | 0.42  (0.31, 0.56) | 0.47  (0.36, 0.60) | 0.53  (0.42, 0.65) | 0.58  (0.48, 0.70) | 0.63  (0.54, 0.74) | 0.69  (0.60, 0.79) | 0.74  (0.65, 0.84) | 0.84  (0.73, 0.95) | 0.91  (0.79, 1.04) | 0.95  (0.81, 1.11) | 1.00  (0.82, 1.20) |
| **NH** | 1.08  (0.65, 1.70) | 1.06  (0.68, 1.58) | 1.02  (0.70, 1.45) | 0.99  (0.72, 1.34) | 0.94  (0.72, 1.22) | 0.90  (0.71, 1.12) | 0.85  (0.69, 1.03) | 0.83  (0.68, 1.01) | 0.79  (0.64, 0.97) | 0.73  (0.57, 0.92) | 0.67  (0.50, 0.88) |
| **NJ** | 0.34  (0.26, 0.44) | 0.40  (0.32, 0.49) | 0.45  (0.37, 0.55) | 0.52  (0.44, 0.61) | 0.58  (0.50, 0.66) | 0.65  (0.58, 0.73) | 0.72  (0.65, 0.79) | 0.83  (0.75, 0.91) | 0.93  (0.83, 1.03) | 1.00  (0.88, 1.13) | 1.07  (0.92, 1.24) |
| **NM** | 0.35  (0.25, 0.48) | 0.40  (0.30, 0.53) | 0.46  (0.36, 0.59) | 0.53  (0.43, 0.66) | 0.60  (0.50, 0.72) | 0.68  (0.58, 0.79) | 0.76  (0.66, 0.86) | 0.88  (0.77, 1.00) | 0.99  (0.86, 1.14) | 1.08  (0.92, 1.26) | 1.17  (0.96, 1.41) |
| **NV** | 0.52  (0.27, 0.92) | 0.50  (0.28, 0.85) | 0.48  (0.28, 0.78) | 0.46  (0.28, 0.72) | 0.43  (0.26, 0.67) | 0.41  (0.25, 0.64) | 0.38  (0.22, 0.60) | 0.37  (0.21, 0.61) | 0.35  (0.19, 0.60) | 0.32  (0.16, 0.57) | 0.29  (0.13, 0.55) |
| **NY** | 0.80  (0.67, 0.94) | 0.83  (0.72, 0.96) | 0.86  (0.76, 0.98) | 0.89  (0.80, 1.00) | 0.90  (0.82, 0.99) | 0.92  (0.84, 1.00) | 0.92  (0.85, 1.00) | 0.96  (0.88, 1.05) | 0.98  (0.89, 1.07) | 0.95  (0.85, 1.06) | 0.93  (0.82, 1.05) |
| **OH** | 0.51  (0.37, 0.67) | 0.52  (0.40, 0.67) | 0.53  (0.43, 0.66) | 0.54  (0.45, 0.65) | 0.54  (0.46, 0.63) | 0.54  (0.47, 0.62) | 0.53  (0.46, 0.61) | 0.55  (0.47, 0.64) | 0.55  (0.46, 0.65) | 0.53  (0.43, 0.64) | 0.51  (0.40, 0.64) |
| **OK** | 0.80  (0.33, 1.64) | 0.80  (0.35, 1.59) | 0.80  (0.37, 1.55) | 0.81  (0.38, 1.53) | 0.80  (0.37, 1.50) | 0.79  (0.36, 1.50) | 0.77  (0.35, 1.49) | 0.79  (0.34, 1.57) | 0.79  (0.32, 1.62) | 0.75  (0.29, 1.62) | 0.73  (0.26, 1.64) |
| **OR** | 0.64  (0.47, 0.87) | 0.66  (0.50, 0.86) | 0.68  (0.53, 0.85) | 0.69  (0.56, 0.84) | 0.69  (0.57, 0.82) | 0.69  (0.58, 0.82) | 0.68  (0.57, 0.82) | 0.71  (0.58, 0.86) | 0.71  (0.56, 0.88) | 0.68  (0.52, 0.88) | 0.66  (0.48, 0.88) |
| **PA** | 0.62  (0.53, 0.72) | 0.66  (0.58, 0.74) | 0.69  (0.62, 0.77) | 0.72  (0.66, 0.80) | 0.74  (0.69, 0.81) | 0.77  (0.71, 0.83) | 0.78  (0.73, 0.84) | 0.83  (0.77, 0.89) | 0.85  (0.78, 0.92) | 0.84  (0.77, 0.92) | 0.83  (0.75, 0.93) |
| **RI** | 11.04  (6.74, 17.09) | 10.67  (6.94, 15.70) | 10.22  (7.01, 14.39) | 9.81  (7.01, 13.32) | 9.21  (6.78, 12.22) | 8.70  (6.47, 11.46) | 8.12  (5.97, 10.79) | 7.90  (5.63, 10.76) | 7.46  (5.09, 10.54) | 6.79  (4.38, 10.01) | 6.20  (3.75, 9.59) |
| **SC** | 1.22  (1.01, 1.46) | 1.23  (1.05, 1.43) | 1.23  (1.07, 1.40) | 1.23  (1.09, 1.38) | 1.20  (1.09, 1.33) | 1.18  (1.07, 1.30) | 1.15  (1.04, 1.26) | 1.16  (1.04, 1.28) | 1.13  (1.00, 1.28) | 1.07  (0.92, 1.23) | 1.01  (0.85, 1.18) |
| **SD** | 0.86  (0.44, 1.52) | 0.94  (0.51, 1.57) | 1.01  (0.59, 1.61) | 1.08  (0.67, 1.66) | 1.14  (0.74, 1.68) | 1.21  (0.81, 1.74) | 1.27  (0.87, 1.80) | 1.39  (0.94, 1.98) | 1.48  (0.98, 2.14) | 1.51  (0.97, 2.26) | 1.55  (0.94, 2.41) |
| **TN** | 0.68  (0.41, 1.05) | 0.69  (0.44, 1.02) | 0.69  (0.48, 0.98) | 0.70  (0.51, 0.94) | 0.69  (0.53, 0.88) | 0.69  (0.56, 0.84) | 0.67  (0.57, 0.79) | 0.69  (0.60, 0.79) | 0.68  (0.59, 0.78) | 0.65  (0.55, 0.77) | 0.63  (0.51, 0.76) |
| **TX** | 0.56  (0.47, 0.67) | 0.58  (0.50, 0.67) | 0.59  (0.52, 0.67) | 0.61  (0.54, 0.68) | 0.61  (0.55, 0.67) | 0.61  (0.56, 0.66) | 0.60  (0.55, 0.66) | 0.62  (0.56, 0.68) | 0.62  (0.56, 0.69) | 0.60  (0.53, 0.67) | 0.58  (0.50, 0.66) |
| **UT** | 0.20  (0.09, 0.38) | 0.20  (0.09, 0.36) | 0.20  (0.10, 0.35) | 0.20  (0.10, 0.34) | 0.19  (0.10, 0.33) | 0.19  (0.10, 0.33) | 0.19  (0.10, 0.32) | 0.19  (0.10, 0.34) | 0.19  (0.09, 0.35) | 0.18  (0.08, 0.34) | 0.17  (0.07, 0.35) |
| **VA** | 1.45  (1.21, 1.73) | 1.36  (1.16, 1.58) | 1.26  (1.10, 1.43) | 1.16  (1.04, 1.30) | 1.05  (0.95, 1.16) | 0.96  (0.88, 1.04) | 0.86  (0.79, 0.93) | 0.80  (0.73, 0.88) | 0.73  (0.65, 0.81) | 0.63  (0.56, 0.71) | 0.55  (0.48, 0.64) |
| **VT** | 0.52  (0.33, 0.78) | 0.56  (0.37, 0.80) | 0.59  (0.42, 0.82) | 0.64  (0.47, 0.84) | 0.66  (0.51, 0.85) | 0.70  (0.55, 0.88) | 0.73  (0.57, 0.91) | 0.79  (0.61, 0.99) | 0.82  (0.63, 1.07) | 0.83  (0.61, 1.12) | 0.85  (0.59, 1.18) |
| **WA** | 0.51  (0.31, 0.80) | 0.51  (0.33, 0.76) | 0.51  (0.34, 0.72) | 0.50  (0.35, 0.69) | 0.49  (0.35, 0.66) | 0.47  (0.34, 0.64) | 0.46  (0.32, 0.63) | 0.46  (0.31, 0.65) | 0.45  (0.29, 0.66) | 0.42  (0.26, 0.65) | 0.40  (0.23, 0.64) |
| **WI** | 0.72  (0.53, 0.96) | 0.74  (0.57, 0.95) | 0.76  (0.60, 0.94) | 0.77  (0.64, 0.93) | 0.77  (0.65, 0.91) | 0.78  (0.67, 0.89) | 0.77  (0.67, 0.87) | 0.79  (0.70, 0.90) | 0.79  (0.69, 0.91) | 0.76  (0.65, 0.89) | 0.74  (0.61, 0.88) |
| **WV** | 0.66  (0.50, 0.85) | 0.69  (0.54, 0.86) | 0.71  (0.58, 0.86) | 0.74  (0.62, 0.87) | 0.75  (0.65, 0.86) | 0.76  (0.68, 0.86) | 0.77  (0.69, 0.86) | 0.81  (0.72, 0.90) | 0.82  (0.72, 0.92) | 0.80  (0.69, 0.92) | 0.78  (0.66, 0.92) |
| **WY** | 0.62  (0.37, 0.97) | 0.64  (0.41, 0.95) | 0.65  (0.44, 0.93) | 0.67  (0.47, 0.91) | 0.67  (0.49, 0.88) | 0.67  (0.51, 0.87) | 0.67  (0.51, 0.86) | 0.69  (0.53, 0.89) | 0.70  (0.52, 0.91) | 0.68  (0.49, 0.91) | 0.66  (0.45, 0.92) |

Note: Models are adjusted for age group, sex, race and ethnicity; expected case counts calculated via indirect standardization were used as the reference for each state and year

**Table S3: Adjusted prevalence ratio for aspergillosis post-(2020-2023) compared to pre- (2013-2019) emergence of COVID-19 by demographic group–United States**

| **Ethnicity, Residence Type, Race** | **Adjusted Prevalence Ratio** | **95% CI** | ***P*-value** |
| --- | --- | --- | --- |
| Hispanic or Latino |  |  |  |
| *Rural* |  |  |  |
| American Indian/Alaska Native | 1.47 | 1.07‚ 2.02 | 0.02 |
| Asian/Pacific Islander | 1.63 | 1.37‚ 1.95 | <0.001 |
| Black or African American | 1.33 | 1.16‚ 1.53 | <0.001 |
| Other | 1.27 | 1.14‚ 1.42 | <0.001 |
| White | 1.27 | 1.14‚ 1.42 | <0.001 |
| *Urban* |  |  |  |
| American Indian/Alaska Native | 1.39 | 1.01‚ 1.90 | 0.04 |
| Asian/Pacific Islander | 1.54 | 1.32‚ 1.81 | <0.001 |
| Black or African American | 1.25 | 1.11‚ 1.42 | <0.001 |
| Other | 1.20 | 1.10‚ 1.31 | <0.001 |
| White | 1.20 | 1.10‚ 1.31 | <0.001 |
| Non-Hispanic or Latino |  |  |  |
| *Rural* |  |  |  |
| American Indian/Alaska Native | 1.24 | 0.91‚ 1.69 | 0.17 |
| Asian/Pacific Islander | 1.38 | 1.19‚ 1.61 | <0.001 |
| Black or African American | 1.13 | 1.01‚ 1.26 | 0.04 |
| Other | 1.08 | 0.96‚ 1.21 | 0.22 |
| White | 1.08 | 1.00‚ 1.16 | 0.06 |
| *Urban* |  |  |  |
| American Indian/Alaska Native | 1.17 | 0.86‚ 1.59 | 0.31 |
| Asian/Pacific Islander | 1.31 | 1.14‚ 1.50 | <0.001 |
| Black or African American | 1.06 | 0.97‚ 1.17 | 0.20 |
| Other | 1.02 | 0.92‚ 1.12 | 0.76 |
| White | 1.02 | 0.96‚ 1.07 | 0.55 |

**Table S4: Trends in COVID-19 associated changes in aspergillosis prevalence: regression results of adjusted prevalence ratios by race and ethnicity–United States, 2013-2023**

| **Variable** | **Estimate** | **Standard Error** | ***P*-value** |
| --- | --- | --- | --- |
| (Intercept) | 1.04 | 0.03 | <0.001 |
| White | *ref* | *ref* | *ref* |
| American Indian or Alaska Native | 0.18 | 0.03 | <0.001 |
| Asian/Pacific Islander | 0.32 | 0.03 | <0.001 |
| Black or African American | 0.05 | 0.03 | 0.14 |
| Some Other Race | -0.001 | 0.03 | 0.97 |
| Hispanic or Latino^*^ | 0.21 | 0.02 | <0.001 |

*^Note: ref = reference group,^* ^*^*^Hispanic or Latino is compared to non-Hispanic or Latino^*

**Equation S1: Post-Stratification Standardization Weights**

$$w_{i}=\frac{{Population Propotion}_{i}}{{Sample Proportion}_{i}}$$

$$w_{i}=\frac{P_{pop}\left( Age=i \right)\times P_{pop}\left( Race/Ethnicity=i \right)\times P_{pop}\left( Gender=i \right)\times N_{pop}}{P_{sample}\left( Age=i \right)\times P_{sample}\left( Race/Ethnicity=i \right)\times P_{sample}\left( Gender=i \right)\times N_{sample}}$$

Where:

$w_{i}$ = calculated per state and year

$P_{pop}$ = probability in the population data source

$P_{sample}$ = probability in the sample data source

**Figure S1: Total aspergillosis diagnoses (unweighted raw counts) by state in Oracle EHR Real World Data–United States, 2013-2023**
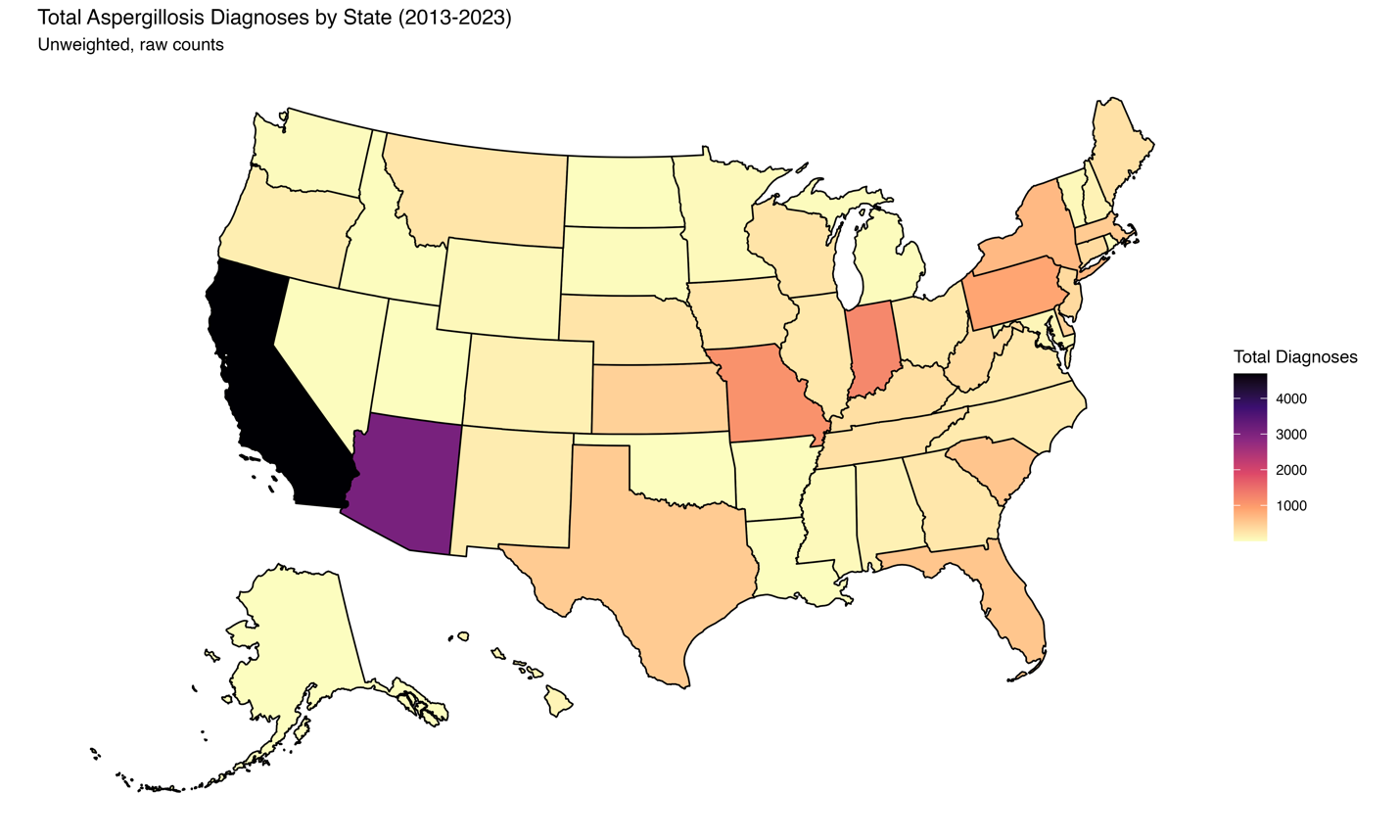

**Figure S2: Population-based (weighted) prevalence of aspergillosis diagnoses per 100,000 person-years in Oracle EHR Real World Data, by year–United States, 2013-2023**

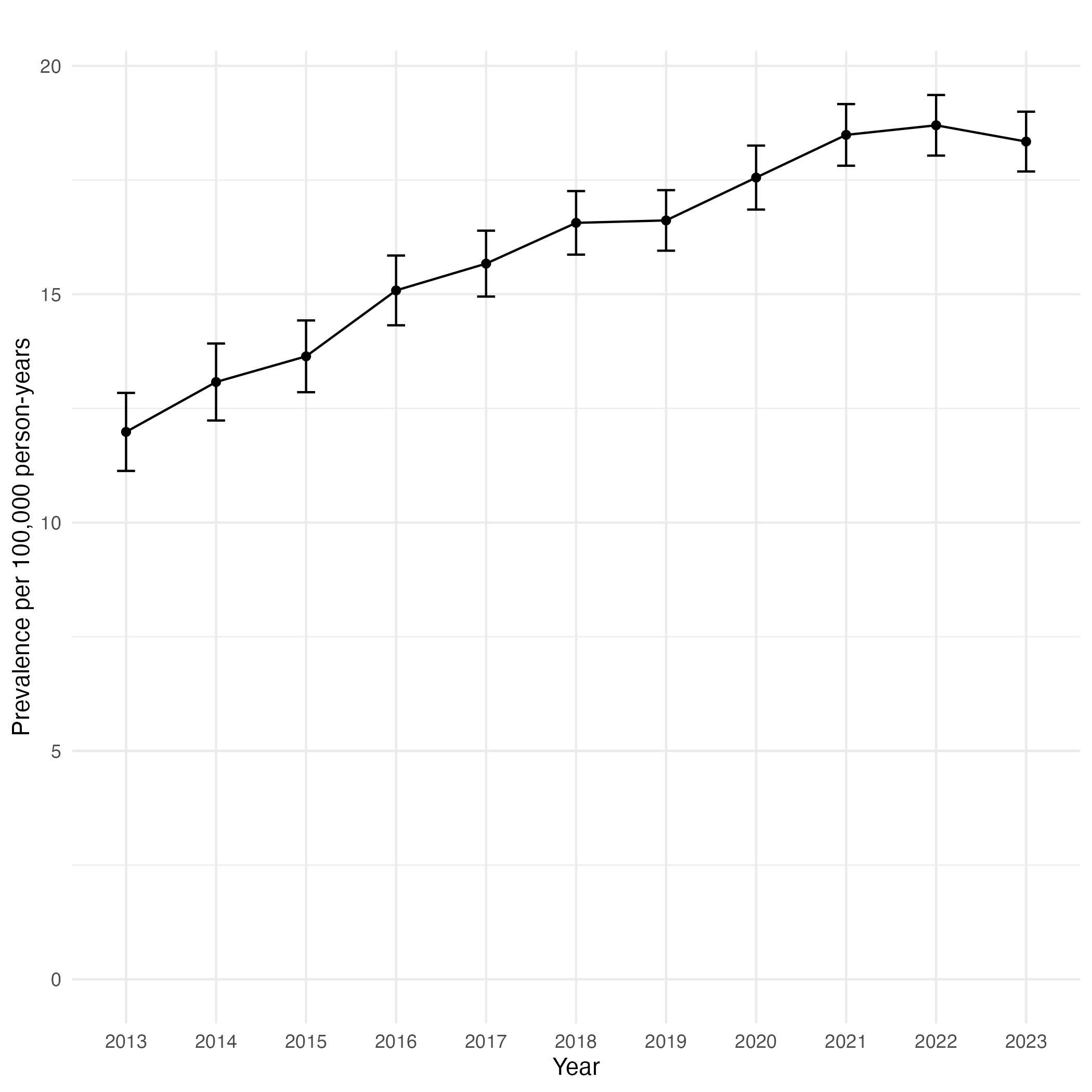
